# Single-dose effectiveness of mpox vaccine in Quebec, Canada: test-negative design with and without adjustment for self-reported exposure risk

**DOI:** 10.1101/2023.07.09.23292425

**Authors:** Nicholas Brousseau, Sara Carazo, Yossi Febriani, Lauriane Padet, Sandrine Hegg-Deloye, Geneviève Cadieux, Geneviève Bergeron, Judith Fafard, Hugues Charest, Gilles Lambert, Denis Talbot, Jean Longtin, Alexandre Dumont-Blais, Steve Bastien, Virginie Dalpé, Pierre-Henri Minot, Gaston De Serres, Danuta M. Skowronski

## Abstract

**Introduction:** During the 2022 mpox outbreak, the province of Quebec, Canada prioritized first doses for pre-exposure vaccination of people at high mpox risk, delaying second doses due to limited supply. We estimated single-dose mpox vaccine effectiveness (VE) adjusting for virus exposure risk based only on surrogate indicators available within administrative databases (e.g. clinical record of sexually transmitted infections), or supplemented by self-reported risk factor information (e.g. sexual contacts).

**Methods:** We conducted a test-negative case-control study between June 19 and September 24, 2022. Information from administrative databases was supplemented by a questionnaire collecting self-reported risk factors specific to the three-week period before testing. Two study populations were assessed: all within the administrative databases (All-Admin) and the subset completing the questionnaire (Sub-Quest). Logistic regression models were adjusted for age, calendar-time and exposure-risk, the latter based on administrative indicators only (All-Admin and Sub-Quest populations) or supplemented by questionnaire information (Sub-Quest population).

**Results:** There were 532 All-Admin participants, of which 199 (37%) belonged to Sub-Quest. With exposure-risk adjustment based only on administrative indicators, single-dose mpox VE among All-Admin and Sub-Quest populations was similar at 35% (95%CI:-2-59) and 30% (95%CI:-38-64), respectively. With adjustment supplemented by questionnaire information, Sub-Quest VE increased to 65% (95%CI:1-87). Protection against severe outcomes was higher (VE=82%; 95%CI:-50-98) but with overlapping confidence intervals.

**Conclusions:** One vaccine dose reduced mpox risk by about two-thirds when adjustment incorporated self-reported risk factors, but by only one-third when adjustment relied solely upon administrative indicators. Inadequate adjustment for exposure risk may substantially under-estimate mpox VE.

## Introduction

In May 2022, an mpox (formerly known as monkeypox) outbreak started in several non-endemic countries prompting the World Health Organization (WHO) to declare a public health emergency of international concern on July 23, 2022[1]. In the city of Montréal (Province of Qubec, Canada) the first mpox cases were laboratory-confirmed on May 20, 2022. As of June 1, 2023, 79% of the 529 cases reported in Quebec were among Montreal residents. Transmission mostly occurred among communities of gay, bisexual, trans, queer (GBTQ) men and other men who have sex with men (gbMSM). Outbreak control primarily relied on surveillance, case and contact management, and pre-exposure vaccination with the Modified Vaccinia Ankara—Bavarian Nordic [MVA-BN] vaccine (Imvamune [Canada], Jynneos [USA], Imvanex [Europe]).

MVA-BN is a third-generation non-replicating vaccine, approved by Health Canada in 2020 to prevent *orthopoxvirus* infections[2]. Post-exposure MVA-BN vaccination began in Quebec on May 30, 2022 and on June 3, 2022, Quebec became one of the first jurisdictions to offer pre-exposure vaccination[3,4], with gradual roll-out first targeting gbMSM at high-risk of exposure, and progressively extended to sex workers and people working in sex-on-premise venues. The standard MVA-BN vaccination regimen consists of two subcutaneous doses, 28 days apart. Due to limited supply, providing the first MVA-BN dose to as many as possible, including to foreign visitors, was prioritized in Quebec[5] and Canada[6], thereby delaying second doses. In Quebec, the second dose was initially restricted to immunocompromised persons and became offered widely as of October 6, 2022. As of June 1, 2023, a total of 31,000 first doses and 13,000 second doses had been administered in Quebec.

The efficacy of MVA-BN pre-exposure vaccination against mpox has never been demonstrated in clinical trials. Several observational studies have attempted to evaluate single-dose vaccine effectiveness (VE) against mpox disease with widely varying estimates ranging between 36%[7] and 86%[8]. Most studies, however, have relied upon administrative databases lacking specific details on exposure risk, and using surrogate indicators as proxy for mpox exposure risk. With limited vaccine doses targeted toward high-risk individuals, differential exposure risk between vaccinated and unvaccinated persons may pose a particular threat to valid VE estimation.

In this test-negative case-control study, we estimated the effectiveness of subcutaneous single-dose MVA-BN vaccination against symptomatic mpox infection, including moderate-to-severe disease. We compared mpox VE estimates with adjustment for exposure risk based solely on surrogate indicators of risk, as typically available within administrative databases, or supplemented by self-reported risk factor information specific to the three-week period before mpox testing.

## Methods

### Design

We conducted a test-negative case-control study using skin or mucous membrane specimens that were tested for *orthopoxvirus* infections by nucleic acid test (NAT) at the Quebec Public Health Laboratory (QPHL). Cases were test-positive. Controls were individuals with only negative NAT tests. Among those with multiple test-negative specimens, only the first-ever specimen for orthopoxvirus testing, collected during the study period, was used.

### Data sources

#### Administrative data

We obtained information on cases and controls from several administrative and surveillance databases. The QPHL data included all *orthopoxviruses* NAT test results, date and place of specimen collection, age, sex and human immunodeficiency virus (HIV) status (any past positive test result). Mpox vaccination data were retrieved from the Provincial Immunization Registry, a populational registry where all immunizations have to be recorded by law[9]. Information on sexually transmitted infection (STI) testing (chlamydia, gonorrhea and syphilis) was retrieved from the *Dossier Santé Québec* database containing individual clinical information, while positive STI tests were identified within the provincial notifiable disease registry.

For cases, mpox clinical data (hospitalization, complications) were also obtained from the provincial notifiable disease registry. Tecovirimat treatment information was retrieved from the only designated pharmacy allowed to distribute the medication in the province (*Centre hospitalier universitaire de Montréal*).

#### Questionnaire data

We supplemented administrative data with self-reported sociodemographic and exposure data collected from both cases and controls using a standardized questionnaire available in French, English and Spanish. The questionnaire was adapted from tools used for public health epidemiological investigations (**Supplementary Methods**). Between January 10 and March 27, 2023, cases and controls were invited by email to complete the questionnaire on-line, with three reminders. Trained interviewers attempted to contact non-responders and those without an e-mail address by phone, with three reminders. Participation was actively promoted by the main health and STI prevention organization serving gbMSM communities in Montreal (RÉZO)[10], and $30 financial compensation was given to participants.

The questionnaire collected vaccination status and additional demographic and clinical information such as ethnocultural background, immunosuppression, and HIV pre-exposure prophylaxis (PrEP). When MVA-BN vaccination was not in the Immunization Registry but reported by participants with a specific date, the participant was considered vaccinated. The questionnaire specifically queried details related to mpox exposure risk during the 3 weeks before specimen collection including contact with a person with mpox symptoms, number of sexual partners, drug use before or during sexual activity, sex involving > 2 people, attending sex-on-premise venues, and skin-to-skin contacts during a festive event.

### Study population

The chosen study period spanned June 19 to September 24, 2022 (epidemiological weeks 26-38), taking into account vaccine roll-out, a 14-day lag to mount effective single-dose immunity, and epidemic evolution with cases peaking during June and July, and subsiding by mid-September 2022 (**Figure 1**).

**Figure 1.**
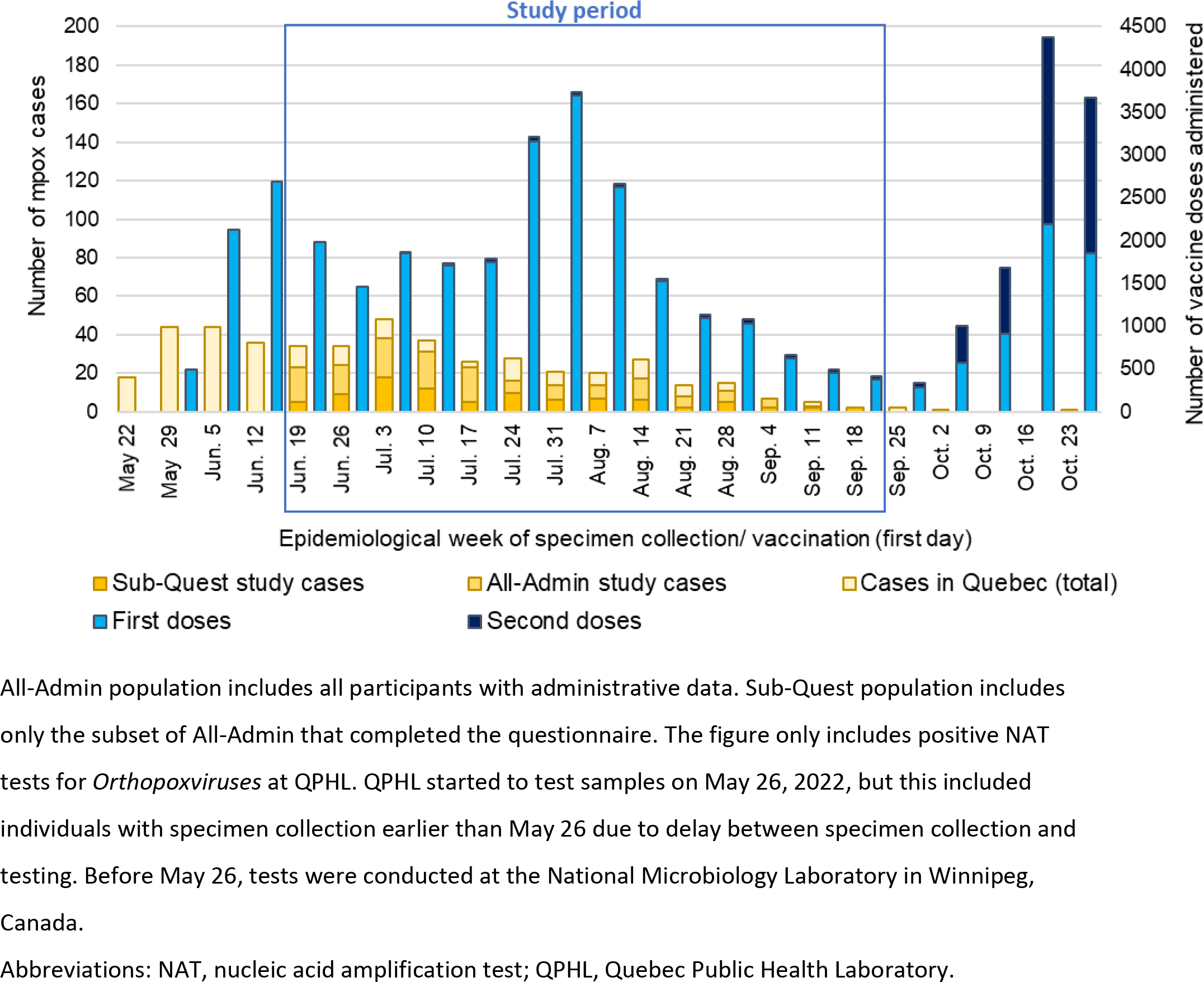
Weekly distribution of study cases and vaccine administration in the province of Quebec, May 22 to October 29, 2022.

Eligible specimens were restricted to those collected in Montreal where most cases occurred. Since most cases were men (according to health administrative databases), we excluded women. We also excluded individuals aged <18 years, those who tested positive before the study period, those whose only specimens were blood, serum or cerebrospinal fluid samples (not indicative of a skin or mucous membrane lesion), asymptomatic individuals, and persons residing outside of the province.

Two study populations were analyzed: (1) all participants meeting eligibility criteria and using administrative data only (All-Admin); and (2) the subset of All-Admin participants who completed the questionnaire (Sub-Quest).

### Vaccination status

Vaccinated status was defined by receipt of a single dose of MVA-BN vaccine ≥14 days before specimen collection. Those who had not received any MVA-BN vaccine dose before specimen collection were considered unvaccinated. We excluded individuals who had received one MVA-BN dose <14 days or two doses at any time before specimen collection.

### Outcomes

Single-dose VE was primarily assessed against any laboratory-confirmed symptomatic infection, among the All-Admin population and the Sub-Quest population separately. We also estimated VE against moderate-to-severe disease defined by mpox disease-related hospitalization, having had a complication or having received tecovirimat treatment. VE against moderate-to-severe disease could only be undertaken using the All-Admin population owing to sample size considerations. The same control group of any symptomatic but test-negative individuals was used in assessing VE for both clinical outcomes.

### Statistical analysis

Logistic regression models compared the adjusted odds of being a case versus a control by vaccine status. VE was derived as (1 – odds ratio [OR_adjusted_]) x 100. Analysis was performed using SAS (version 9.4).

For the All-Admin population, we adjusted models for the following variables available from linked administrative databases: age group, calendar time, surrogate indicator of exposure risk ≤6 months before the mpox test (no test for STI, only negative STI test results, ≥ 1 positive STI test results), and HIV status (see **Figure 3** for categories used).

For the Sub-Quest population, we compared VE based on two adjustment approaches: (1) adjusting as per above for variables available in administrative databases and (2) adjusting for variables available in administrative databases and also for information additionally available from the questionnaire: ethnocultural background, immunosuppression, HIV status/PrEP use, contact with a person who has mpox symptoms, number of sexual partners, drug use before or during sexual activity, sex involving > 2 people, attending sex-on-premise venues, skin-to-skin contacts during a festive event (see **Figure 3** for categories used). We defaulted to the questionnaire responses for HIV status where discordant from administrative databases. There were no missing data for any variable included in the model.

To assess possible recall bias, we compared the number of sexual partners from the notifiable disease registry recorded during public health investigation of reported cases (Summer 2022) versus questionnaire capture (Winter 2023). We also undertook four additional sensitivity analyses of VE within the Sub-Quest population, including: (1) extension of the study period to include epidemiological weeks 22-24 and 39-44; (2) vaccination defined as ≥ 21-days before specimen collection; (3) restriction to younger adults 18-49 years old; and (4) restriction to participants with self-reported PrEP use or HIV diagnosis.

### Ethics

In the context of a public health emergency, this work was conducted under the legal mandate provided by the Quebec Public Health Act which allowed access to relevant administrative data[9] and was also approved by the *CHU de Québec-Université Laval* ethics committee.

## Results

### Study sample

From June 19 to September 24 2022, there were 1025 individuals with a specimen submitted from Montreal for mpox NAT analysis (277 cases; 748 controls) (**Figure 2**). Among these, 448 (44%) were excluded with main reasons being: asymptomatic and tested for a research project (348/448; 78%), and woman (81/448; 18%). After applying all exclusion criteria, 532 individuals (231 cases; 301 controls) were included in the All-Admin analysis. The Sub-Quest subset who answered the questionnaire included 199 (37%) participants (91 cases; 108 controls).

**Figure 2.**
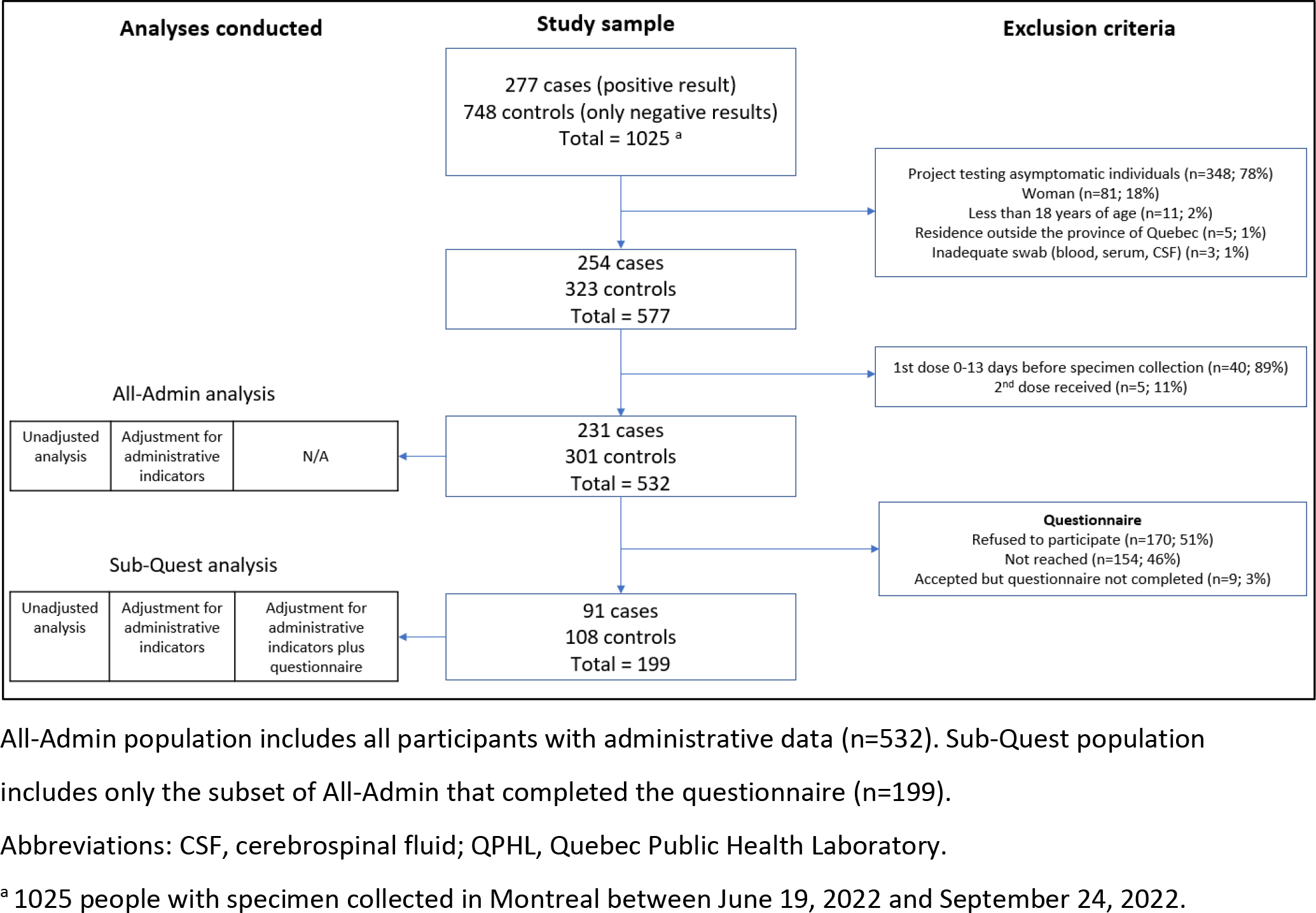
Flowchart of study population.

### Case and control description

Among the 532 All-Admin participants, half (116/231; 50%) NAT-positive cases were reported before mid-July whereas most (162/301; 54%) NAT-negative controls were reported later in August and September (**Table 1**). Slightly fewer cases than controls were vaccinated (26% vs. 34%, respectively). Few cases or controls were ≥50 years old (16% vs 22%). More cases than controls were HIV-positive (22% vs. 14%) or had a positive STI test in the 6 months before mpox testing (26% vs. 10%).

**Table 1.** Characteristics of cases and controls among All-Admin and Sub-Quest populations, June 19 to September 24, 2022.

|  | All-Admin population<br>(n=532) |  | Sub-Quest population<br>(n=199) |  |
| --- | --- | --- | --- | --- |
|  | Cases<br>N (%) | Controls<br>N (%) | Cases<br>N (%) | Controls<br>N (%) |
| **Overall** | 231 | 301 | 91 | 108 |
| **ADMINISTRATIVE DATABASE VARIABLES** |  |  |  |  |
| **Age (years)** |  |  |  |  |
| 18-29 | 50 (22) | 90 (30) | 21 (23) | 28 (26) |
| 30-49 | 143 (62) | 146 (49) | 57 (63) | 55 (51) |
| 50+ | 38 (16) | 65 (22) | 13 (14) | 25 (23) |
| **Epidemiological week of specimen collection** |  |  |  |  |
| 25-28 (June 19 - July 16) | 116 (50) | 68 (23) | 44 (48) | 26 (24) |
| 29-31 (July 17 - August 6) | 53 (23) | 71 (24) | 21 (23) | 26 (24) |
| 32-34 (August 7 - August 27) | 39 (17) | 90 (30) | 15 (16) | 27 (25) |
| 35-38 (August 28 - September 24) | 23 (10) | 72 (24) | 11 (12) | 29 (27) |
| **Days between symptom onset and testing (median; IQR)** | 6 (4 - 9)<sup>a</sup> | NA | 6 (4 - 8)<sup>a</sup> | NA |
| **Vaccination status** |  |  |  |  |
| Unvaccinated | 172 (74) | 199 (66) | 57 (66) | 55 (51) |
| 1 dose ≥14 days before testing | 59 (26) | 102 (34) | 34 (37) | 53 (49) |
| 1 dose ≥21 days before testing | 52 (23) | 94 (31) | 30 (33) | 47 (47) |
| **HIV infection (QPHL data)** | 51 (22) | 42 (14) | 15 (16) | 13 (12) |
| **Test done for chlamydia OR gonorrhea<sup>b</sup>** | 188 (81) | 199 (66) | 76 (84) | 81 (75) |
| **Test done for syphilis<sup>b</sup>** | 183 (79) | 202 (67) | 74 (81) | 78 (72) |
| **Positive test for chlamydia<sup>b</sup>** | 36 (16) | 11 (4) | 16 (18) | <5 (<5) |
| **Positive test for gonorrhea<sup>b</sup>** | 39 (17) | 20 (7) | 17 (19) | 11 (10) |
| **Positive test for primary syphilis<sup>b</sup>** | <5 (<5) | <5 (<5) | <5 (<5) | <5 (<5) |
| **Overall: tests for STI<sup>b</sup>** |  |  |  |  |
| No test for STI | 39 (17) | 90 (30) | 13 (14) | 24 (22) |
| ≥ 1 test for STI, no positive results | 132 (57) | 181 (60) | 53 (58) | 71 (66) |
| ≥ 1 positive STI test results | 60 (26) | 30 (10) | 25 (27) | 13 (12) |
| **QUESTIONNAIRE-ELICITED VARIABLES** |  |  |  |  |
| **Gender** |  |  |  |  |
| Man | NA | NA | 90 (99) | 103 (95) |
| Other (woman, non-binary, PNA) | NA | NA | <5 (<5) | 5 (5) |
| **Ethnocultural background** |  |  |  |  |
| French Canadian | NA | NA | 40 (44) | 56 (52) |
| English Canadian | NA | NA | 5 (5) | 8 (7) |
| Other, do not know, PNA | NA | NA | 46 (51) | 44 (41) |
| **Immunosuppression (excluding HIV)** | NA | NA | 7 (8) | <5 (<5) |
| **HIV infection/PrEP use (study questionnaire)** |  |  |  |  |
| HIV infection | NA | NA | 21 (23) | 15 (14) |
| No HIV infection, PrEP use | NA | NA | 43 (47) | 36 (33) |
| No HIV infection, no PrEP | NA | NA | 27 (30) | 57 (53) |
| **Skin-to-skin contact with person with mpox symptoms<sup>c</sup>** | NA | NA | 39 (43) | 14 (13) |
| **Number of sexual partners<sup>c</sup>** |  |  |  |  |
| None | NA | NA | 10 (11) | 27 (25) |
| Only regular partner(s) | NA | NA | 9 (10) | 15 (14) |
| 1-2 non-regular partners | NA | NA | 28 (31) | 27 (25) |
| 3-4 non-regular partners | NA | NA | 11 (12) | 20 (19) |
| 5 or more non-regular partners | NA | NA | 17 (19) | 10 (9) |
| Do not know, PNA | NA | NA | 16 (18) | 9 (8) |
| **Use of drugs before/during sexual activity<sup>c</sup>** | NA | NA | 32 (35) | 35 (32) |
| **Sex involving > 2 people<sup>c</sup>** | NA | NA | 29 (32) | 25 (23) |
| **Attended sex-on-premise venues<sup>c</sup>** | NA | NA | 36 (40) | 29 (27) |
| **Skin-to-skin contact (with or without sex) during a festive event<sup>c</sup>** | NA | NA | 34 (37) | 36 (33) |
| **Overall: at least one exposure<sup>c</sup>** | NA | NA | 88 (97) | 93 (86) |

All-Admin population includes all participants with administrative data. Sub-Quest population includes only the subset of All-Admin that completed the questionnaire. Cells sizes <5 hidden for privacy reasons. Abbreviations: HIV, human immunodeficiency virus; IQR, interquartile range; NA, not applicable; PNA, prefers not to answer; PrEP, pre-exposure prophylaxis; QPHL, Quebec Public Health Laboratory; STI, sexually transmitted infection. <sup>a</sup> Twenty-one missing data in the All-Admin population; four missing data in the Sub-Quest population. <sup>b</sup> ≤ 6 months before mpox specimen collection. <sup>c</sup> All self-reported exposures within 3 weeks of mpox specimen collection.

Results based on administrative data were similar between the All-admin and Sub-Quest participants (**Table 1**). By questionnaire, cases reported greater mpox exposure risk than controls during the 3 weeks before specimen collection including ≥5 non-regular sexual partners (19% vs. 9%, respectively) and sex involving > 2 people (32% vs. 23%). Nearly all cases (88/91; 97%) but fewer controls (93/108; 86%) had at least one self-reported mpox exposure risk factor during the 3 weeks before testing. More cases were HIV-negative but taking PrEP (47%) compared to controls (33%). Our questionnaire appeared slightly more sensitive to identify HIV-positive individuals than administrative databases, as 18% of Sub-Quest participants self-reported an HIV infection, but 14% of Sub-Quest participants had HIV according to administrative (QPHL) data.

In the questionnaire (completed on average 26 weeks post-diagnosis), 10/91 (10%) Sub-Quest cases reported no sexual partners in the 3 weeks before specimen collection. In contrast, the notifiable disease registry (data collected on average two weeks post-diagnosis) indicated that all 91 Sub-Quest cases had reported sexual contact(s) during that period. There was otherwise good concordance between these two data sources with respect to the number of sexual partners reported (**Supplementary Table 1**).

### Single-dose MVA-BN vaccine effectiveness

Single-dose MVA-BN VE estimates against symptomatic infection among All-Admin and Sub-Quest populations were 35% (95%CI: −2-59) and 30% (95%CI: −38-64), respectively, with adjustment for age, calendar time and exposure risk based only upon administrative indicators (**Figure 3, Supplementary Table 2**).

**Figure 3.**
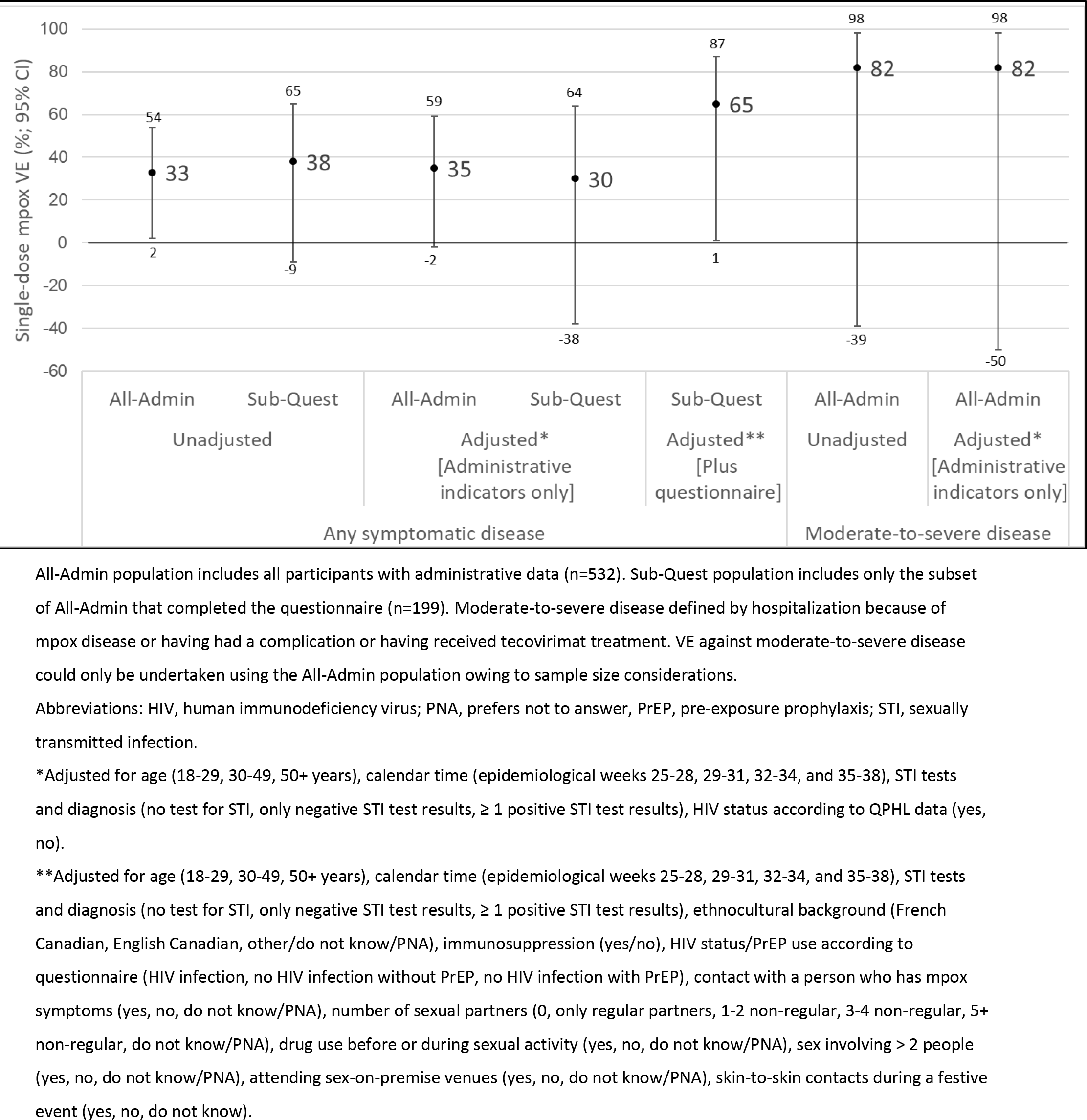
Single-dose MVA-BN vaccine effectiveness against mpox infection by study population (All-Admin or Sub-Quest), exposure risk adjustment (administrative indicators only or supplemented by questionnaire) and clinical profile (any symptomatic or moderate-severe disease)

Single-dose MVA-BN VE within the Sub-Quest population increased to 65% (95%CI:1-87) when adjusting for age, calendar time and exposure risk with the latter further supplemented by questionnaire information (**Figure 3, Supplementary Table 2**). The most influential questionnaire variable was HIV PrEP use, with Sub-Quest VE of 50% (95%CI:-8-77) when only that questionnaire variable supplemented age, calendar time and administrative indicators of exposure risk. However, other questionnaire covariates (without adjusting for HIV PreP) were also influential with VE of 45% (95%CI:-37-78) when supplemental to age, calendar time and administrative indicators of exposure risk.

There were 12/208 (6%) moderate-to-severe cases during the study period within the All-Admin population (of whom just 3 responded to the study questionnaire). Of these 12, three were hospitalized, six (two also hospitalized) experienced complications, and five received tecovirimat treatment (one also hospitalized). Only 1/12 moderate-to-severe cases (8%) had received MVA-BN vaccine compared with 101/287 controls (35%) suggesting higher VE against moderate-to-severe disease (82%; 95%CI: −39, 98) (Figure 3, Supplementary Table 2).

### Sensitivity analyses

Sensitivity analyses were performed on the Sub-Quest population, inclusive of questionnaire covariates. When compared with the primary analysis (VE=65%; 95%CI:1-87), VE point estimates were slightly lower, between 50-63%, when extending the study period earlier from May 29 or later to November 5, based on vaccination ≥21 days before specimen collection or when restricting to people with an HIV diagnosis or taking PrEP. VE was higher at 75% (95%CI:23-92) with restriction to adults 18-49 years (**Table 2**).

**Table 2.**
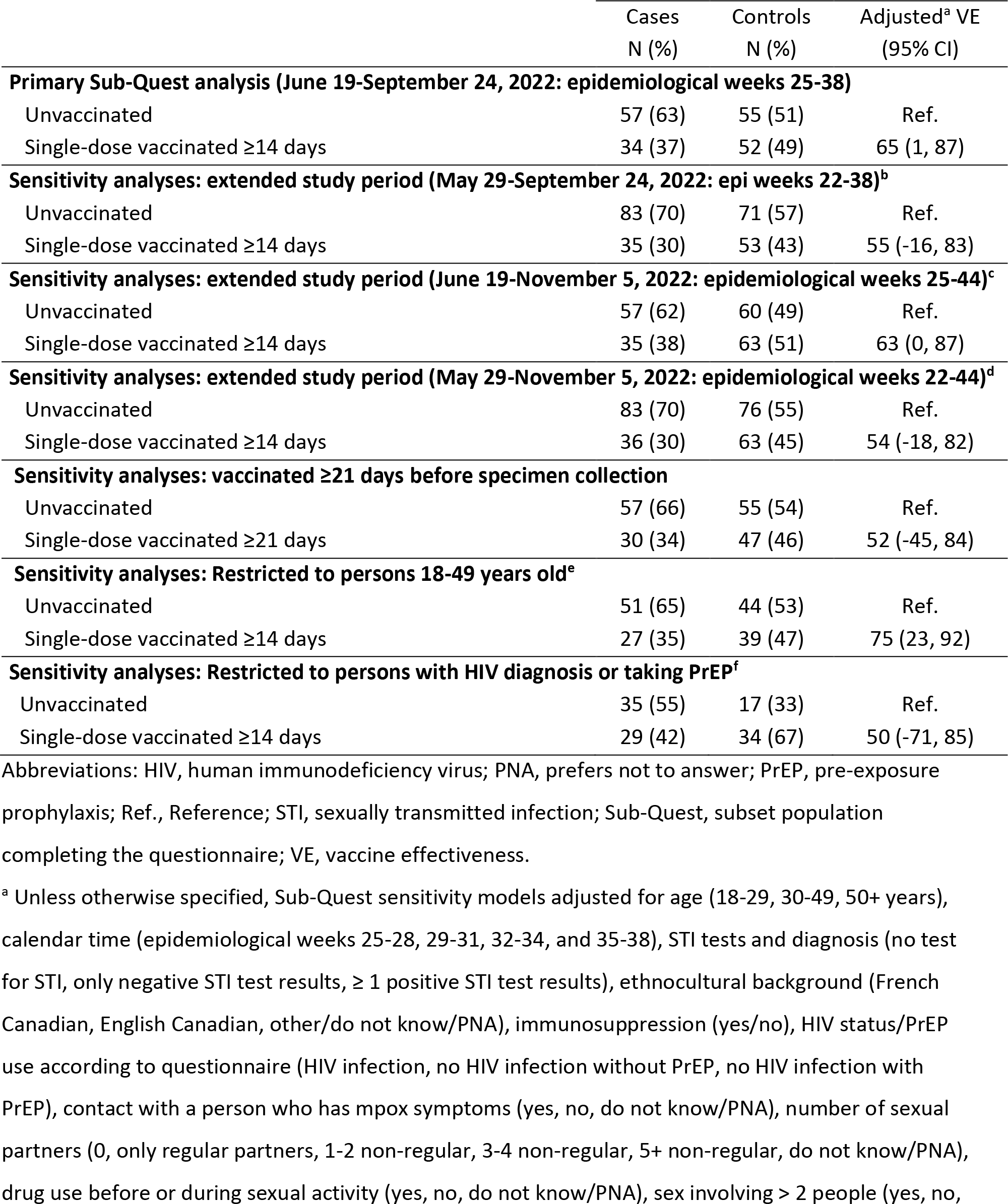
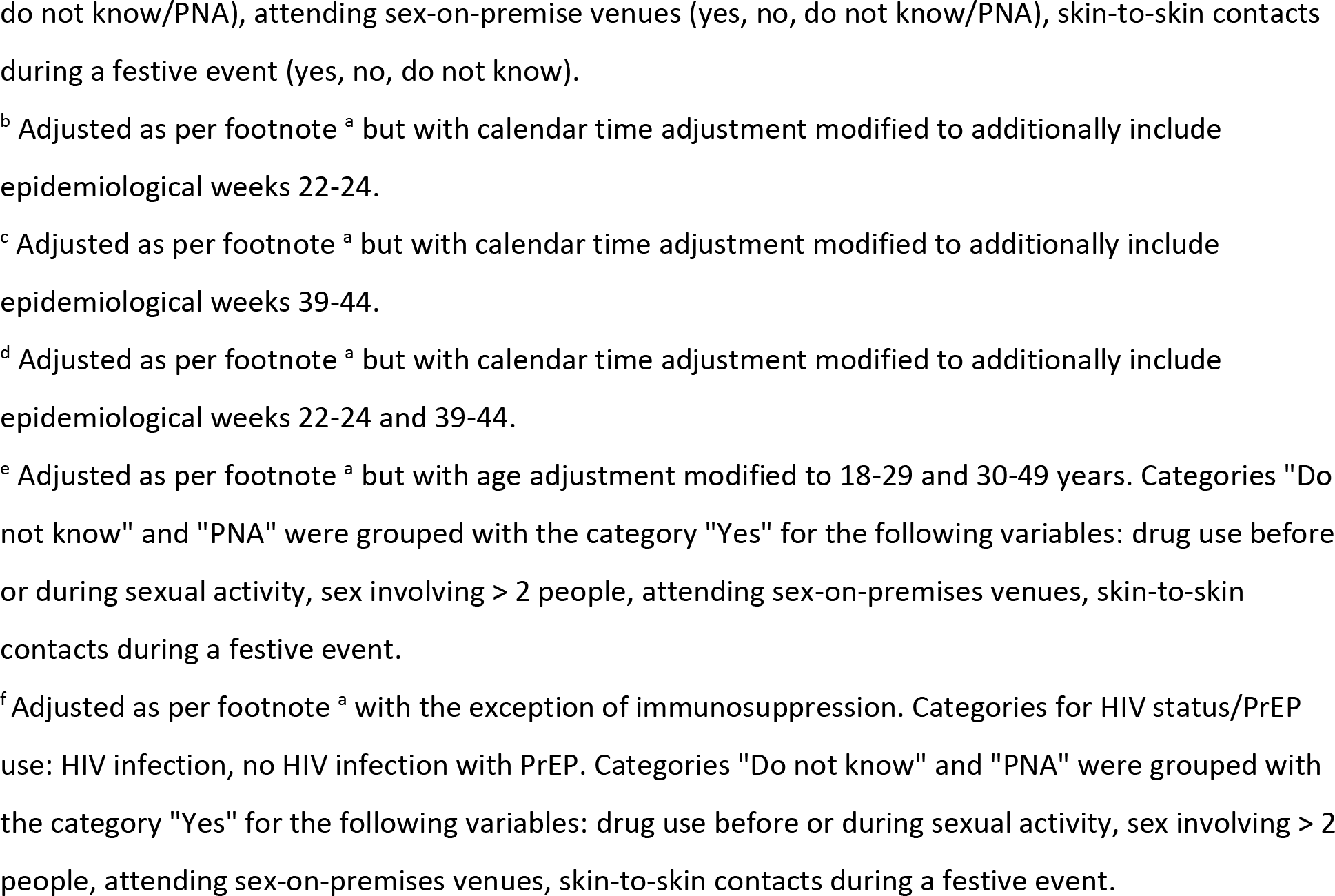
Sensitivity analyses of single-dose MVA-BN vaccine effectiveness against symptomatic mpox infection, Sub-Quest population inclusive of questionnaire covariates.

## Discussion

One MVA-BN vaccine dose during an mpox outbreak affecting gbMSM communities in Montreal was associated with about two-thirds reduction in the risk of symptomatic mpox infection when adjustment for mpox exposure risk included self-reported risk factors. The estimated risk reduction was much lower (roughly one-third) when adjustment used only surrogate indicators of exposure risk available in administrative databases. In sensitivity analyses, VE point estimates ranged 50-75%, indicating a 2-4-fold lower mpox risk among single-dose vaccinated compared to unvaccinated individuals. Recognizing the limited sample size, our findings also suggest greater protection against moderate-to-severe disease compared to milder disease with estimated VE of about 80% corresponding to 5-fold risk reduction.

Prior MVA-BN VE studies based upon other observational designs including case-control, case-coverage and cohort approaches have reported widely ranging single-dose VE against mpox, with estimates spanning 36%[7] to ≥78%[7,8,11,12]. These analyses, however, were limited by the use of aggregate data only[11] and/or failure to adequately adjust for confounders such as differential exposure risk by vaccine status[7,8] and/or calendar time[8,11,12]. In addition to exposure risk adjustment, calendar-time adjustment is also important to consider in observational estimation of VE, as previously well-established for influenza and COVID-19. When the incidence of mpox decreases with time across the study period and vaccinated person-time accrues later than unvaccinated person-time, VE will be over-estimated[13]. Other mpox VE investigations based on classical matched case-control designs included controls that may have not presented with mpox symptoms, with single-dose mpox VE estimates of 68%[14] and 75%[15]. These estimates are slightly higher than our own, which was bases instead on a test-negative design that simultaneously standardized for healthcare seeking and testing indication, in addition to controlling for exposure risk and calendar time.

The lower single-dose VE estimates previously reported by Deputy et al.[7] (36%) is similar to our single-dose VE estimate among our All-Admin (35%) and Sub-Quest (30%) participants before incorporating self-reported risk factors into covariate adjustment within the Sub-Quest population. Of note, the comparability between our All-Admin and Sub-Quest populations when adjusting for the same covariates is partially reassuring against selection bias potentially influencing the subset completing the study questionnaire. The Deputy et al. results were based on a large case-control study applied to a nationwide electronic health record database, matching cases and controls ≥18 years with incident diagnosis of HIV infection or an order for PrEP based upon week of the index event. In our study, additionally adjusting for HIV PrEP alone had the greatest impact on VE (50%), but adjustment for other self-reported risk factors further increased our overall single-dose VE estimate (65%). This insight was achieved through actively contacting and querying cases and controls to document mpox exposure risk, an added step not typically undertaken for controls. When we restricted our Sub-Quest population to people taking PrEP or with an HIV diagnosis, our single-dose VE estimate (50%) was still higher, but closer to that of Deputy et al.[7] That further stratification, however, was accompanied by reduced sample size and wide confidence intervals. More research is needed to determine if restricting to people taking HIV PrEP or adjusting for this variable as a surrogate indicator within administrative databases could support valid VE estimation if/when self-reported mpox exposure risk cannot otherwise be adequately documented[16].

Our finding of higher single-dose VE against moderate-to-severe disease was based on a single hospitalized vaccinated case, lacking statistical power but nevertheless consistent with immunogenicity studies. While two MVA-BN doses in non-primed individuals induced low levels of poorly neutralizing antibody[17], a single dose induced robust T-cell responses[18,19], with the latter more strongly correlated with protection against severe outcomes in general (e.g. against COVID-19 disease[20]).

Other indirect evidence includes a cases series of mpox cases (without controls) in the United States by Farrar et al.[21] that could not derive VE estimates, but found a lower proportion of 1-dose vaccinated (2.1%) versus unvaccinated (7.5%) cases that were hospitalized. While severe mpox fortunately remains infrequent, more definitive VE evaluation against that outcome will require much larger sample size.

This study has several limitations. The main limitation was the small sample size attributable to the contained outbreak and participation rate to the questionnaire which limited our capacity to undertake further stratified analyses (e.g., to assess waning with time). Although we standardized for being symptomatic at the time of testing, controls may not have had symptoms of similar severity to cases[22]. We can expect some level of recall bias with our questionnaire, as there was imperfect agreement between information (e.g. sexual contacts) collected on average within two-weeks (case investigation) versus 26 weeks (study questionnaire) after the diagnosis. Our questionnaire still enabled us to adjust for self-reported mpox exposure risk, including variables less likely to be affected by recall bias, such as HIV PrEP. We cannot rule out residual confounding due to unmeasured factors. Finally, we provide evidence for single-dose protection delivered through subcutaneous injection but cannot comment on other injection route or schedules (e.g. intradermal or two-dose). Other studies that have evaluated two doses show improved VE[7,11,14,15], underscoring that a complete series remains preferable when supply is sufficient.

In conclusion, during the 2022 mpox outbreak in Quebec, single-dose MVA-BN vaccine recipients were at 2-4-fold lower risk of symptomatic mpox disease compared to unvaccinated individuals and vaccine protection may be higher against severe outcomes. Incomplete adjustment for mpox exposure risk, details of which are generally not available in administrative databases, may substantially under-estimate mpox VE. In the context of MVA-BN scarcity and substantial single-dose protection, prioritizing first doses and delaying second dose delivery can maximise benefit to the targeted population.

## Data Availability

Data produced in the present study are available upon request.

## Acknowledgements

We first want to thank all participants who answered the questionnaire. We also thank RÉZO community, an organization for health and STIs prevention aimed at GBTQ men and other MSM, for their help and expertise support throughout the project, and ensuring that the project would be well received by involved communities and answer their questions. We also thank Paul Le Guerrier and Robert Allard for their collaboration on the study design. We finally thank Eveline Toth, Hélène Venables and Annick Des Cormiers for their collaboration, and Christiane Audet, Charles Bellavance, Lucie Ferland, Sophie Grenier, Gabrielle Guilbault, Sarah Shakibaian, and Maud Vallée for their help with data collection and data extraction.

All authors participated in research design. N.B, S.C, L.P., S.H.G., J.F., and H.C. participated in the acquisition of the data. N.B, S.C, Y.F., S.H.G., D.T., G.D.S., and D.M.S. contributed to data analysis. N.B. wrote the first draft of the manuscript and all authors participated in the interpretation of data and reviewed the article.

