## Supplementary materials for "Single-dose effectiveness of mpox vaccine in Quebec, Canada: test-negative design with and without adjustment for self-reported exposure risk"

### Supplementary methods

Questionnaire

**Do you identify as (more than one choice possible)?**
Male
Female
Non-binary
I prefer to describe myself as: ________________
I prefer not to respond

**Which of the following best describes your ethnocultural background?**
African (other than North Africa)
East, Southeast or South Asian
Indigenous (First Nations, Inuit, Métis)
Canadian English
French Canadian
Caribbean
West European, East European
Latin American
Middle Eastern, Arabic, North African
Mixed ethnic origin, please specify: ____________________
I use another term to describe my ethnocultural background, please specify: ___________
I don't know
I prefer not to respond

**In Canada, until 1971, many people were vaccinated against smallpox (against human smallpox, not mpox). The vaccine was administered in the shoulder and left a scar on the skin. To the best of your knowledge, have you received the HUMAN smallpox vaccine?**
Yes
No
I don't know
I prefer not to answer

**According to the Quebec vaccination registry which we had access to as part of this project, here is the information we have regarding the mpox (monkeypox) vaccine doses you have received since 2022:**
**Number of mpox vaccine (Imvamune) doses received: (NvaccinVS)**
**Date of the 1st dose: (dateVSdose1)**
**Date of the 2nd dose: (dateVSdose2)**
**Is this information correct?**

Yes
No
I don’t know
***If No,***
**Please provide the correct information:**
Number of doses of vaccine (Imvamune) received against mpox: ___________________
Date of the 1st dose if applicable): _______________________
Date of the 2nd dose (if applicable): __________________________

**The mpox vaccine (Imvamune) is sometimes recommended after recent contact (less than 14 days) with someone who has mpox. Did you receive the vaccine for this reason?**
No (I received the vaccine before being exposed)
Yes (I received the vaccine after being exposed, less than 14 days after contact with someone who has mpox)
I don’t know
I prefer not to respond

**The following categories are examples of health conditions that can affect the immune system (defence system against infections). Please check all boxes that apply to you.**
Active cancer
Chemotherapy or active radiation therapy (treatment for cancer)
Recent organ transplant
Taking steroids like prednisone or cortisone on a regular basis. Please specify: _____________________
Taking medicine to treat autoimmune diseases such as arthritis, lupus, ulcerative colitis or Crohn's disease. Please specify: ___________________________
HIV infection
Other, please specify: _______________
I don't know if I have one of these health conditions
I don’t have any if these health conditions
I prefer not to respond
***If no HIV infection,***
**Do you take PrEP (HIV pre-exposure prophylaxis)?**
Yes
No
I don’t know
I prefer not to respond

**To your knowledge, have you been diagnosed with mpox (did you have mpox)?**
Yes
No
I don’t know
I prefer not to respond

**In the 3 weeks before your test for mpox (including the day of your test) (datetest), did you have any lesions (sores, redness) on the skin or on the mucous membranes of the mouth, anus or genitals?**

Yes

No

I don't know

I prefer not to respond

**Mpox is transmitted primarily through prolonged and/or intense skin-to-skin contact. In the 3 weeks before your test for mpox, (datetest), did you have this type of contact with someone who was soon after diagnosed with mpox?**

Yes

No

I don't know

I prefer not to respond

**Mpox is transmitted primarily through prolonged and/or intense skin-to-skin contact. In the 3 weeks before your test for mpox (datetest), did you have this type of contact with someone who had mpox symptoms, but was not diagnosed with mpox?**

Yes

No

I don't know

I prefer not to respond

**In the 3 weeks before your test for mpox (datetest), did you have one or more sexual partners?**

Yes

No

I don't know

I prefer not to respond.

***If yes,***

If yes, how many (even if approximate)? ______

I don't know

I prefer not to respond

***If participant provides a number***

Of these, how many (even if approximate) were not regular partners? _______

I don't know

I prefer not to respond

**In the 3 weeks before your test for mpox (datetest), did you use any of the following drugs before or during sexual activity (Tina, Meth, Speed (Amphetamine), Poppers (Amyl nitrite), Juice, GH (GHB), K, Ket (GHB), Cocaine)?**

Yes, I have taken at least one of these drugs
No, I have not taken any of these drugs
I don’t know
I prefer not to respond

**In the 3 weeks before your test for mpox (datetest), have you had sex involving more than two people at the same time?**

Yes
No
I don’t know
I prefer not to respond

**In the 3 weeks before your test for mpox (datetest), have you attended one or more places with sex on premises (e.g.: private or public festive evening, sauna, park, etc.)?**

Yes
No
I don’t know
I prefer not to respond

**In the 3 weeks before your test for mpox (datetest), do you remember having close, prolonged skin-to-skin contact (with or without sex) at a festive event?**

Yes
No
I don’t know
I prefer not to respond

**Supplementary Table 1. Comparison of self-reported exposure ≤3 weeks before positive test during as elicited by study questionnaire versus public health case investigation**

| | **Study questionnaire**[a] | **Public health case investigation**[b] |
|---|---|---|
| | N (%) | N (%) |
| **Number of sexual partners** | | |
| 0 | 10 (11%) | 0 (0%) |
| 1-2 | 27 (30%) | 33 (36%) |
| 3-4 | 10 (11%) | 11 (12%) |
| 5+ | 25 (27%) | 23 (25%) |
| No answer | 19 (21%) | 24 (26%) |

[a] Information obtained by study questionnaire on average 26 weeks after specimen collection
[b] Information obtained within the notifiable disease registry, collected through public health case investigation on average 2 weeks after specimen collection

**Supplementary Table 2. Single-dose MVA-BN vaccine effectiveness against mpox infection by study population (All-Admin or Sub-Quest), exposure risk adjustment (administrative indicators only or supplemented by questionnaire) and clinical profile (any symptomatic or moderate-severe disease), June 19 to September 24, 2022**

| | VE (%) | 95%CI |
|---|---|---|
| **Any symptomatic disease** | | |
| Unadjusted Analysis | | |
| All-Admin participants | 33 | (2, 54) |
| Sub-Quest participants | 38 | (-9, 65) |
| Adjusted analysis (administrative indicators only) [a] | | |
| All-Admin participants | 35 | (-2, 59) |
| Sub-Quest participants | 30 | (-38, 64) |
| Adjusted analysis (administrative indicators plus questionnaire) [b] | | |
| Sub-Quest participants | 65 | (1, 87) |
| **Moderate-to-severe disease** | | |
| Unadjusted Analysis | | |
| All-Admin participants | 82 | (-39, 98) |
| Adjusted analysis (administrative indicators only) [a] | | |
| All-Admin participants | 82 | (-50, 98) |

All-Admin population includes all participants with administrative data (n=532). Sub-Quest population includes only the subset of All-Admin that completed the questionnaire (n=199). Moderate-to-severe disease defined by hospitalization because of mpox disease or having had a complication or having received tecovirimat treatment. VE against moderate-to-severe disease could only be undertaken using the All-Admin population owing to sample size considerations.

Abbreviations: HIV, human immunodeficiency virus; PNA, prefers not to answer, PrEP, pre-exposure prophylaxis; STI, sexually transmitted infection.

[a] Adjusted for age (18-29, 30-49, 50+ years), calendar time (epidemiological weeks 25-28, 29-31, 32-34, and 35-38), STI tests and diagnosis (no test for STI, only negative STI test results, ≥ 1 positive STI test results), HIV status according to QPHL data (yes, no).

[b] Adjusted for age (18-29, 30-49, 50+ years), calendar time (epidemiological weeks 25-28, 29-31, 32-34, and 35-38), STI tests and diagnosis (no test for STI, only negative STI test results, ≥ 1 positive STI test results), ethnocultural background (French Canadian, English Canadian, other/do not know/PNA), immunosuppression (yes/no), HIV status/PrEP use according to questionnaire (HIV infection, no HIV infection without PrEP, no HIV infection with PrEP), contact with a person who has mpox symptoms (yes, no, do not know/PNA), number of sexual partners (0, only regular partners, 1-2 non-regular, 3-4 non-regular, 5+ non-regular, do not know/PNA), drug use before or during sexual activity (yes, no, do not know/PNA), sex involving > 2 people (yes, no, do not know/PNA), attending sex-on-premise venues (yes, no, do not know/PNA), skin-to-skin contacts during a festive event (yes, no, do not know).
